# QiC_3_: A novel automated quantitative immunohistological disease activity index for ileocolonic Crohn’s disease and ulcerative colitis

**DOI:** 10.64898/2026.06.04.26354902

**Authors:** Mohammad Kadivar, Manar Alyamani, Michiko Mori, Maryam Kadivar, Jimmie Jönsson, Erik Hertervig, Olof Grip, Lena Svensson, Jonas S Erjefält, Jan Marsal

## Abstract

**Background:** Histological examination of mucosal tissue in inflammatory bowel diseases (IBD) is a sensitive tool to measure disease activity, and histological remission is emerging as a potentially important treatment target. There are several existing histopathological indices, but they often encompass caveats such as not primarily having been designed to measure the degree of inflammation, encompassing subjective components with poor intra- and interindividual reproducibility, and requiring expert pathologists who are scarce, thus resulting in extended response times.

**Aim:** To construct a new computerized, automated index to objectively measure histological disease activity in the ileal and colonic mucosa, applicable to both Crohn’s disease (CD) and ulcerative colitis (UC).

**Materials and methods:** Ileocolonic biopsies were collected from control subjects and patients with CD or UC. A group of CD patients was sampled before and after 12 weeks of anti-TNFα therapy. Another group of CD and UC patients functioned as a small validation cohort. Epithelial cells, neutrophils, macrophages, and T cells were immunohistochemically stained, followed by digitalization of the color signal and computerized delineation of the epithelial and lamina propria compartments. The various immune cell types within the epithelium and the lamina propria, respectively, were enumerated, and the numbers were compared between control subjects and patients with CD or UC.

**Results:** The numbers of neutrophils and macrophages in the epithelium, and neutrophils in the lamina propria, showed the highest sensitivity and specificity for distinguishing control-subject tissues from CD and UC tissues. These three parameters were thus chosen to construct a new index, named QiC_3_ 1.0, that could separate tissues from control subjects and patients with CD or UC with high precision. It performed equally well in a small validation cohort of patients. The QiC_3_ index correlated well with previously described histopathological indices, fecal calprotectin, and endoscopic scores in UC, but showed worse correlation with endoscopic scores in CD and symptomatic scores. When applying the new index to tissues from CD patients before and after therapy, it showed good responsiveness, demonstrating a distinct amelioration in the microscopic inflammatory status that corresponded well to improvements in histopathological scores.

**Conclusion:** We describe a new quantitative, computerized, automated, non-subjective, and response-sensitive immunohistological index (QiC_3_) for measuring disease activity in ileal and colonic mucosal biopsies, suitable for both CD and UC.

## Introduction

Crohn’s disease (CD) and ulcerative colitis (UC) are the two main forms of inflammatory bowel disease (IBD), and both are chronic and relapsing in character. While UC is confined solely to the mucosa of the large intestine, the inflammatory process in CD may be transmural and affect any part of the entire gastrointestinal tract.^1^ In IBD, disease activity may be measured through various indices evaluating symptoms, quality-of-life, endoscopic appearance, levels of biomarkers, or histopathology; however none of these correlate completely to the others.^2,3^ Symptoms are still the dominating monitoring tool in clinical practice, but the endoscopic appearance is currently the gold standard in judging the level of disease activity.^3,4^ The IBD-field is increasingly embracing a treat-to-target strategy in the management of patients, and endoscopic healing is currently the target of choice.^5^ Voices are being raised, however, that endoscopy may underestimate the extent of disease and that the histopathological picture is more closely linked to clinical prognosis and outcomes.^2,3,6–8^ Histological healing is a more stringent end-point than endoscopic evaluation, but it is less well characterized and less used with regards to defining resolution of inflammation in both clinical practice and trials.^2,3,5,9,10^ This may in part be due to the lack of consensus concerning histological remission in IBD. Furthermore, most indices include parameters that describe tissue architecture which reflect tissue damage rather than the current degree of inflammation.

Researchers and pathologists have developed various histological indices to assess disease activity in UC and CD.^2,3,6,10–12^ However, as a result of the differences in the features of UC and CD, there is no unified histological index to assess disease activity in both forms of IBD.^13^ The Geboes Score (GS)^12^ in UC, and the colonic and ileal Global Histologic Disease Activity Score (GHAS)^11^ in CD have been widely used. GS quantifies architectural changes, chronic and acute inflammatory infiltration, presence of neutrophils in the epithelium, crypt destruction, and presence of erosions or ulcerations; while GHAS is based on epithelial damage, structural alterations, features of acute and chronic inflammatory changes, presence of granuloma, and the number of segmental biopsies affected.^11,12^ Even though it is common to use these indices in histological evaluations, GHAS has not been validated and GS has only been partially validated.^2,3^ GS can either be used by assigning a score from 0-5 depending on the most based on the most severe histopathological element observed in the biopsy, or in a continuous manner assigning a score 0-22 severe sub-item observed in the biopsy, or finally, also in a continuous manner but by summing the scores for each of the 7 items scores 0-3 or 0-4.^14–17^ There is thus a need for a new robust histological index to be used in clinical care, exploratory pre-clinical studies, and clinical trials in IBD. This new index ideally should be global, meaning that it can be used for evaluating inflammation in both ileal and colonic biopsies, and in UC as well as CD. It should also be reliable with high sensitivity and specificity, reproducible, and respond to meaningful alterations in disease severity. Indeed, others have identified the same unmet need, and recently there have been three new indices for IBD suggested: The Robarts histopathological index,^18^ the Nancy index,^19^ and the IBD distribution, chronic features, and activity features (IBD-DCA) score.^10^ However, all of these still require ocular assessments of biopsy sections.

The aim of this study was to explore immunopathological changes in IBD and to use these to develop a non-subjective, computerized, and clinically useful quantitative histological index for measuring the degree of inflammation in ileal and colonic mucosal biopsies from patients with CD or UC. We found that endoscopically inflamed samples from CD and UC patients could be differentiated from samples of control subjects enumerating neutrophils and macrophages confined to the epithelial compartment, and lamina propria (LP) neutrophils. We next formulated a new immunohistological index, QiC_3_, based on these three parameters. The QiC_3_ index was then compared with previously described histological indices for both CD and UC. Its reproducibility was investigated with a small validation cohort of endoscopically active CD and UC samples, and its responsiveness (ability to measure the change in disease activity over time or in response to therapy) was examined in samples from CD patients collected before and after 12 weeks of anti-TNFα treatment.

## Materials and Methods

### Intestinal sampling

Ileocolonic biopsies were collected from control subjects (examined due to iron deficiency or functional gastrointestinal symptoms without any pathological findings on ileocolonoscopy; n=10) and patients with UC (n=14) or CD (n=24). Ten patients with moderate to severe CD (Harvey-Bradshaw Index score of ≥7) were treated with adalimumab (monoclonal anti-TNFα antibody) for 12 weeks (160 mg s.c. at start and 80 mg after two weeks, followed by 40 mg every second week) and sampled before and after treatment. In addition to intestinal biopsies, fecal samples were collected from these ten CD patients. The study was performed in accordance with the declaration of Helsinki and was approved by the regional ethics committees in Lund and Linköping, Sweden (Dnr. 2011/60 and 2011-201-31, respectively). Written informed consent was obtained from all subjects before they were included.

### Fecal calprotectin

Fecal samples were analyzed by enzyme-linked immunosorbent assay (ELISA) according to the manufacturer’s instructions (Bühlmann). Concentration was determined relative to standard curves and expressed as µg/g of feces.

### Endoscopic evaluation

Endoscopic examinations were scored using the Simplified Endoscopic Activity Score for Crohn’s disease (SES-CD)^20^ or the Ulcerative Colitis Endoscopic Index of Severity (UCEIS).^21^

### Histopathological evaluation

Histological and pathological examinations was performed on paraffin-processed biopsy specimens stained with haematoxylin and eosin. The biopsy specimens were read in random order by a single blinded pathologist. For the assessment of histological abnormalities we used two previously described histological scoring system, the continuous (items summed) Geboes Score (cGS) for UC and the colonic and ileal Global Histologic Disease Activity Score (GHAS) for CD.^11,12^ Histologically non-active disease was defined as cGS ≤3 for UC and GHAS ≤2 for CD.

### Immunohistochemistry

A modified double immunohistochemistry protocol was used to quantify CD3, CD68, or myeloperoxidase (MPO) positive cells in the LP or in the epithelium. The immune cell of interest was immune-stained with a dark brown chromogen whereas the epithelial cells in the same section were stained with a green chromogen. Three µm thick paraffin-embedded sections were subjected to heat induced epitope retrieval (HIER) in a pretreatment module (PT-link, DakoCytomation, Denmark) with Tris buffer (pH 6) before immunohistochemical staining at room temperature by an automated immunohistochemistry robot (AutostainerPlus, DakoCytomation / Agilent, Denmark).^22,23^ Sections were blocked with endogenous enzyme block (EnVision™ FLEX Peroxidase-Blocking Reagent, Dako) for 10 minutes and serum free protein block (Dako) for 10 minutes before incubation with primary antibody for 60 minutes. Next, sections were incubated with secondary antibody conjugated with horseradish peroxidase (HRP) labeled polymer (K8010, Dako) for 30 minutes, followed by incubation with diaminobenzidine (DAB) chromogen. After a blocking step with D-block (Biocare Medical, Sweden), the epithelial lining was stained with a cocktail of a mouse anti-human cytokeratin IgG1 antibody (Novocastra, dilution 1:600) and a rabbit anti-β-cathenin (Abcam, ab32572, 1:100). The primary antibodies for immune cell detections were the following: a mouse anti-human CD68 IgG3 antibody (macrophage marker, clone PG-M1, Dado/Agilent, dilution 1:800), rabbit anti-human MPO (neutrophil marker, Dako, dilution 1:7000), and mouse anti-human CD3 (pan T cell marker, Novocastra, dilution 1:400).^23^ Sections were stainied with haematoxylin and placed in a xylene bath before mounting with Pertex mounting medium (Histolab Products, Sweden).

### Image Analysis

After staining the sections were digitalized in an Aperio/Leica Slide Scanner (Scanscope CS, Aperio Technologies, Leica Microsystems, Buffalo Grove, IL, USA operating with a 20x microscope lens) and high-resolution images for all biopsies were, after manual inspection, subjected to automated and unbiased computerized image analysis (Cell Community Viewer, CCV, Medetect AB, Sweden). Briefly, after automated segmentation of background tissue (LP) and the epithelial compartment, the immune cells of interest were detected by color threshold values (compartment segmentation is exemplified in Figure 1). The tissue density for each immune cell type, expressed as the number of pixels occupied by immune cell staining per area, was then automatically calculated for the epithelial compartment and the subepithelial tissue area (LP), respectively. To quantify subepithelially located cells in the LP zone directly underlying the epithelium, the epithelial compartment was expanded by approximately 14 μm, and then the positive pixels in the true epithelial compartment defined in Figure 1C were subtracted from the positive pixels in the expanded epithelial compartment shown in Figure 1D. The number of pixels corresponds directly to the number of positively stained immune cells.

**Figure 1.**
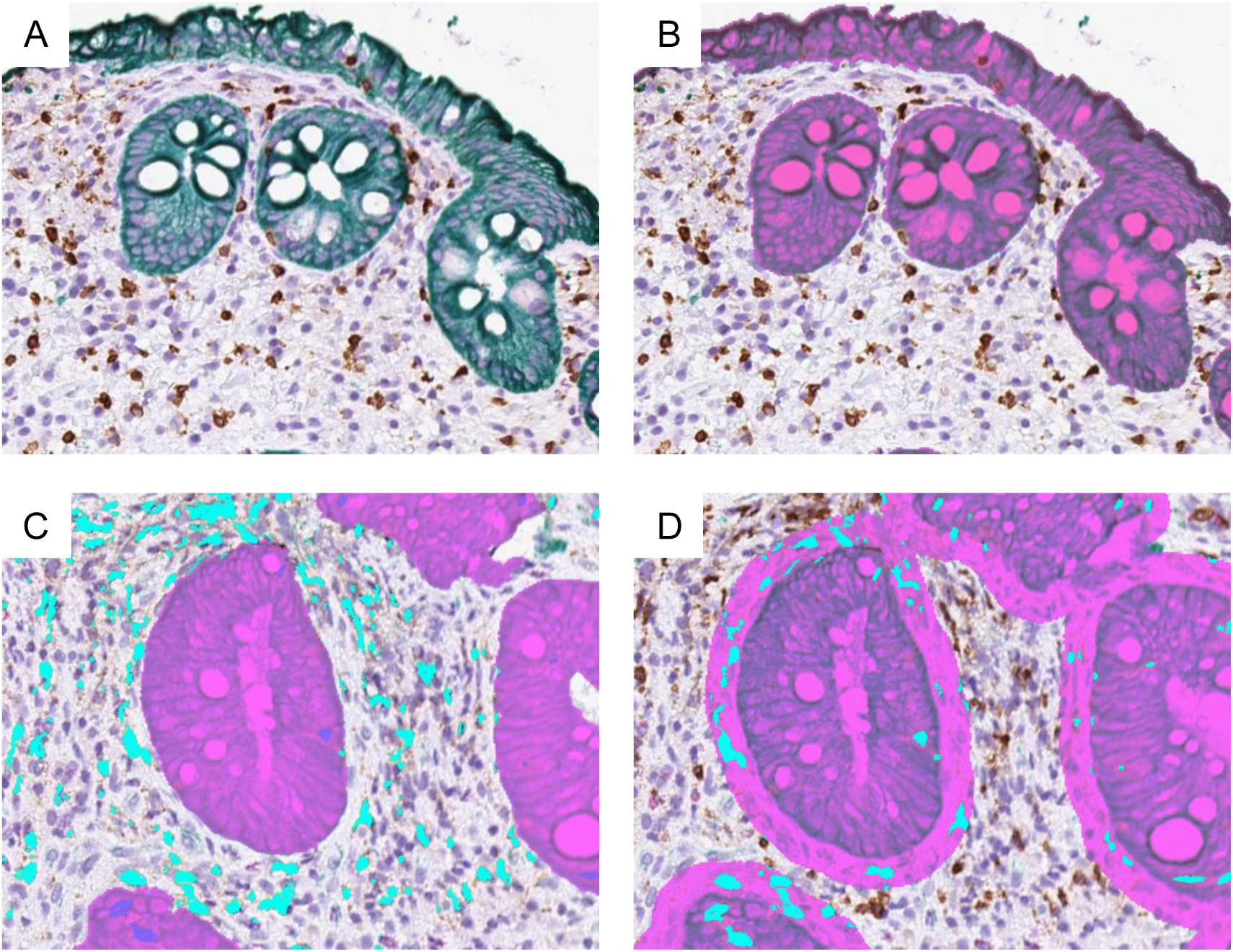
Computerized and automated image analysis for quantification of stained cells in the epithelium and the LP of intestinal tissues. Micrographs illustrate computerized and automated segmentation of intestinal mucosal tissue into epithelium and subepithelial tissue in a colonic biopsy from a UC patient. (A) Epithelial cells were stained for cytokeratin and β-catenin (green) and T cells were stained for CD3 (brown). (B) Computer software detects green pixels as objects, which are used to define the epithelial compartment. Further computerized processing is performed to optimize the compartmentalization, including filling of goblet cell related empty spaces. (C) DAB-stained cells in the LP are identified and labeled light blue, and DAB-stained cells in the epithelial compartment are labeled dark blue. This compartmentalization is then followed by automated quantification of the DAB-stained cells, separately for the LP and the epithelial compartment. (D) Subepithelial cells in the LP zone directly underlying the epithelium were quantified by subtracting the positive pixels in the true epithelial compartment defined in figure C from the positive pixels in the expanded epithelial compartment shown in figure D.

### Statistics

Statistical analyses were performed using GraphPad Prism software v6.0h (GraphPad). Parametric and non-parametric statistical tests were used based on the normal distribution of data (see figure legends for specifications).

### Microarray gene expression analysis for intestinal biopsies

Gut biopsies were collected in RNAlater Stabilization Solution (Ambion, Applied Biosystems) and then frozen on dry ice. Samples were sent on dry ice to the KFB Center of Excellence for Fluorescent Bioanalytics (Regensburg, Germany). Processing of samples for microarray hybridization was carried out at KFB as described in the Affymetrix GeneChip Whole Transcript (WT) Sense Target Labeling Assay manual. Samples were next subjected to the Affymetrix Human Gene 2.1 ST Array and raw data was analyzed using R (version 3.3.0) with Bioconductor package xps. Probe level expression values were normalized and summated using Robust Multiarray Analysis (RMA) algorithm based on the gene model for probesets supported by RefSeq and full-length GenBank transcripts, and with a background correction using Epanechnikov Kernel and only the perfect match probes. Finally, expression data were summarized on the level of human gene symbols and finally normalized by quantile normalization^24^ with the help of R-package limma.

## Results

### Computerized and automated image analysis to quantify immune stained cells in the epithelium and lamina propria of intestinal tissues

To investigate the number of neutrophils, macrophages, and T cells present in the epithelium and the LP, respectively, we analyzed multiple color immunohistochemistry-stained sections from intestinal mucosal biopsies from control subjects and patients with CD or UC (Supplementary Figure 1). The analysis was performed using a computerized and automated image reading system. Gut epithelial cells were stained for β-cathenin and cytokeratin, and based on the digitalized version of this staining, the computer system defined the epithelial compartment (Figure 1). The subepithelial tissue was defined as the LP (Figure 1). The immune cells of interest were stained with another color than the epithelium, followed by digitalization of the color signal.

The computer system defined which immune cells were positively stained, and moreover, in which microanatomical compartment (the epithelium or the LP) the stained cells were situated (Figure 1). Finally, the computer system quantified the number of positive pixels of the different colors, per area, generating data directly corresponding to the number of various immune cells present in the epithelium and the LP, respectively.

### Samples from patients with IBD differed from control samples by the numbers of neutrophils and macrophages in the epithelial compartment and by lamina propria neutrophils

The presence of neutrophils in tissues is a sign of inflammatory activity.^11,12^ Our results demonstrated that the number of neutrophils (MPO^+^ cells) was markedly higher in the epithelial and LP compartments of inflamed intestinal tissues from IBD patients compared to control subjects (Figure 2A). The number of LP neutrophils was higher in the intestinal tissues from CD patients as compared with UC patients, but the difference was not statistically significant (Figure 2A). In addition, we found that the number of macrophages (CD68^+^ cells) in the epithelial compartment was higher in tissues from IBD patients compared to control subjects, while the numbers of macrophages in the LP were similar comparing IBD and control subjects (Figure 2B). To investigate whether the intraepithelial CD68^+^ staining, representing macrophages, potentially was an artifact generated by subepithelial macrophages misinterpreted as being situated intraepithelially, we specifically analyzed the subepithelial zone. However, there was no difference in the numbers of subepithelial macrophages, and thus the differences in CD68^+^ staining were truly intraepithelial. Although there were no statistically significant differences between the numbers of T cells (CD3^+^ cells) in intestinal tissues of IBD patients and control subjects, the mean numbers of LP T cells in CD and UC gut samples were numerically somewhat higher compared with those in control subjects (Figure 2C).

**Figure 2.**
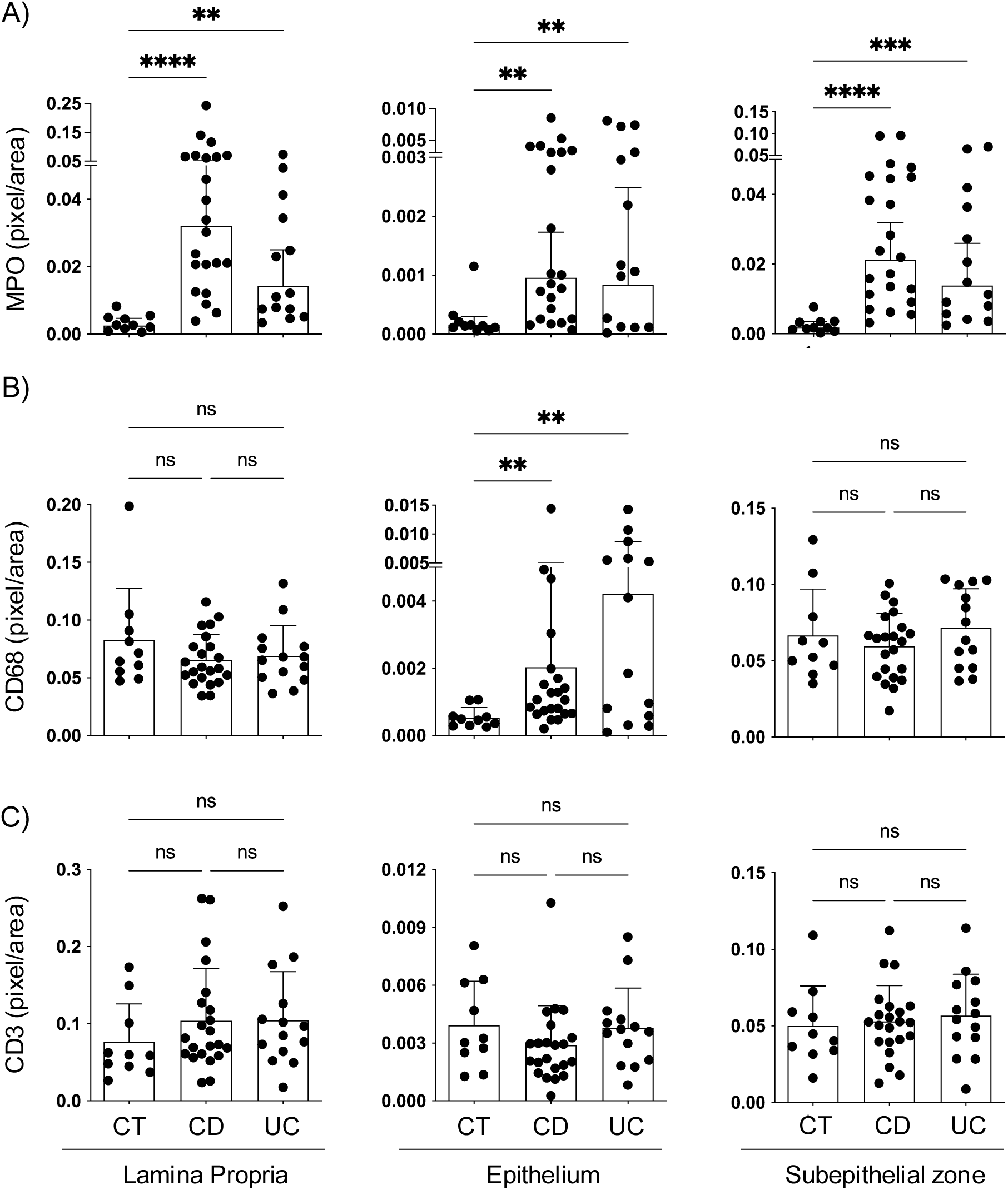
The numbers neutrophils in the LP, the epithelium, and the subepithelial zone, as well as epithelial macrophages, were significantly higher in intestinal tissue samples from IBD patients compared to control subjects. Quantification of (A) neutrophils (MPO^+^ cells), (B) macrophages (CD68^+^ cells), and (C) T cells (CD3^+^ cells), in the LP, the epithelium, and the subepithelial zone, respectively, of control subjects and IBD patients. Each dot represents an individual biopsy and bars are mean±SD. Mann-Whitney test; ns, non significant, *P<0.05, **P<0.001, ***P<0.001.

Next, the ability of each parameter (i.e. numbers of various immune cells in the epithelial and LP compartment, respectively) to discriminate between biopsies from control subjects and IBD patients was investigated using Receiver Operating Characteristic (ROC) analysis (supplementary Figure 2). With this binary classification analysis, we found that epithelial and LP neutrophils, and epithelial macrophages were the parameters with the highest area under the curve (AUC) values, meaning the best ability to differentiate between IBD and control subject intestinal biopsies (supplementary Figure 2). We also showed that the numbers of epithelial and LP neutrophils, and epithelial macrophages correlated to each other, which means that if a section displayed an elevated number for one of these parameters, the probability was high that the numbers for the other parameters would be high too (supplementary Figure 3A). Moreover, the histopathological scores for CD and UC samples showed significant correlations with the numbers of epithelial and LP neutrophils, and epithelial macrophages (supplementary Figure 3B). Therefore, we selected these parameters to be included in the new index.

### Construction of the Quantitative Immunohistochemical Computerized Crohn’s and Colitis (QiC_3_) index

To construct the new index for measuring the degree of inflammatory activity in intestinal mucosal biopsies, we tested to either sum or multiply the parameters that performed the best individually, i.e. the number of epithelial and LP neutrophils, and epithelial macrophages, for controls subject, CD, and UC samples. After summing (Sum-index) or multiplying (Multi-index) these parameters, we transformed the values logarithmically to get normally distributed data (Table 1 and Figure 3). Biopsies from IBD patients that displayed non-active disease as defined by histopathological indices (GHAS ≤2 for CD; cGS ≤3 for UC) and those biopsies displaying active disease were plotted separately. Differences between biopsies with active disease and inactive disease or control subjects, respectively, were statistically significant (Figure 3). According to the ROC analysis of biopsies from control subjects and IBD biopsies with active disease, the Sum-index was more sensitive and specific in terms of differentiating between intestinal biopsies of control subjects and IBD patients than the Multi-index, albeit marginally (Figure 3). We named the Sum-index the Quantitative Immunohistochemical Computerized Crohn’s and Colitis (QiC_3_) 1.0 index which is the term used henceforth. In search for a simplified index, we calculated the MPO positivity (number of neutrophils) only, for the epithelium and the LP collectively, and compared the numbers for IBD biopsies with active disease and the numbers for biopsies from control subjects. The result showed a nearly as good separation as for the QiC_3_ index (supplementary Figure 4).

**Figure 3.**
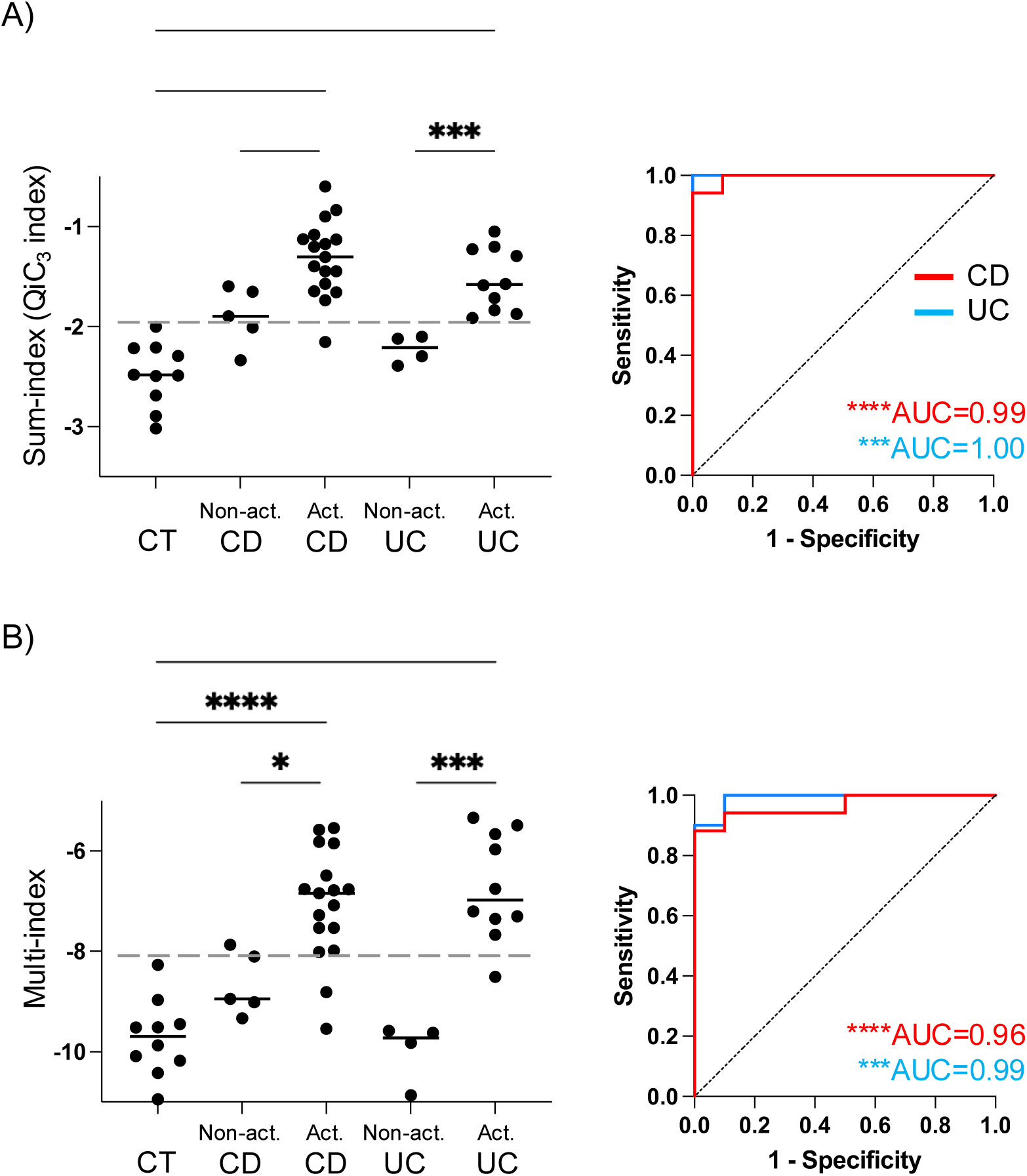
The Sum-index (QiC_3_ index) is the most sensitive and specific candidate scoring system to differentiate between intestinal biopsies of control subjects and biopsies with active inflammation of IBD patients. Graphs show scores generated by the Sum-index (A) and the Multi-index (B) for biopsies from control subjects, and biopsies with active inflammation (Act.) and non-active disease (Non-act.), respectively, of CD and UC patients, respectively. Each dot represents an individual biopsy and bars are mean. Student t-test; ns, non significant, *P<0.05, **P<0.001, ***P<0.001. ROC curves show the sensitivity and specificity of Sum-index (A) and Multi-index (B) for discriminating UC and CD biopsies with active inflammation from biopsies of control subjects. Accuracy was measured by calculations of area under curve (AUC); an AUC of 1.0 represents perfect discrimination. Optimal cutoffs were determined using Youden’s J statistic on ROC curves, with a cutoff of −1.95 for the Sum-index and −8.14 for the Multi-index.

### The absolute numbers of neutrophils, macrophages, and T cells, and QiC_3_ index values were similar for the ileum and the colon

We analyzed the absolute number of epithelial and LP neutrophils, macrophages, and T cells in ileal and colonic biopsies from uninflamed tissues to investigate potential segmental intestinal inherent differences in the QiC_3_ index. Our results demonstrated that the absolute numbers of epithelial and LP neutrophils and T cells, and epithelial macrophages were not significantly different between uninflamed ileum and colon (Figure 4A-C). The exception were LP macrophages that showed slightly higher numbers in the colon (Figure 4B). Indeed, also the QiC_3_ index scores were highly similar for endoscopically uninflamed ileal and colonic samples (Figure 4D).

**Figure 4.**
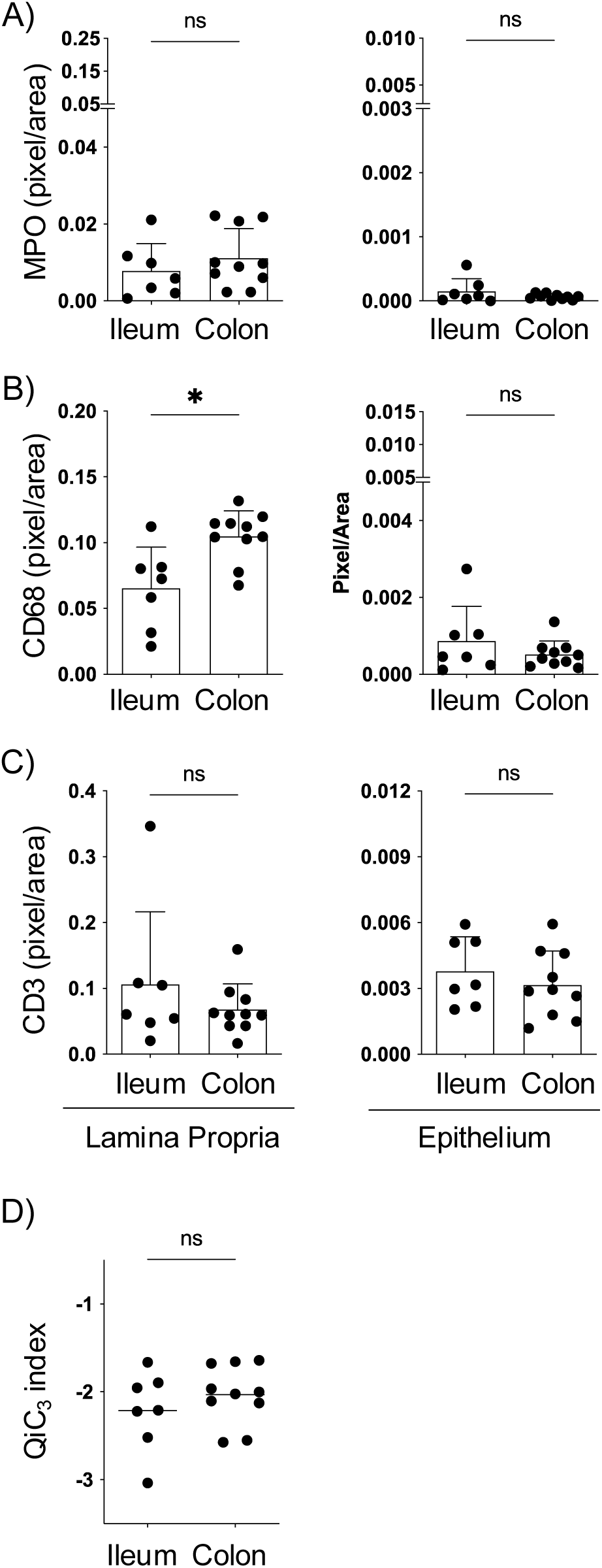
The numbers of neutrophils, macrophages, and T cells in the LP and the epithelium, and the QiC_3_ index, did not differ comparing uninflamed ileal and colonic mucosal samples, except for the numbers of macrophages in the LP. Endoscopically uninflamed ileocolonic mucosa in CD patients was sampled and analyzed. Quantification of (A) neutrophils (MPO^+^ cells), (B) macrophages (CD68^+^ cells), and (C) T cells (CD3^+^ cells) in the LP and the epithelium of uninflamed intestinal biopsies. (D) QiC_3_ index scores for uninflamed ileal and colonic tissue samples. Each dot represents an individual biopsy and bars are mean±SD. Mann-Whitney test; ns, non significant, *P<0.05.

### The QiC_3_ index reflects the histological status of intestinal mucosa

Next, we examined the association of the QiC_3_ index with disease activity evaluated by histological, endoscopic, and symptomatic indices. Our data showed a significant correlation between the QiC_3_ index and histological scores, in both CD and UC patients (Figure 5A). The correlation of the QiC_3_ index and endoscopic scoring (segmental SES-CD and total SES-CD, respectively, for CD and UCEIS for UC) was also statistically significant, with the strongest correlation found in UC (Figure 5B). The correlation between disease activity evaluated by symptoms (scored by HBI and SCCAI, respectively) and the QiC_3_ index was fairly good in CD but less so in UC (Figure 5C).

**Figure 5.**
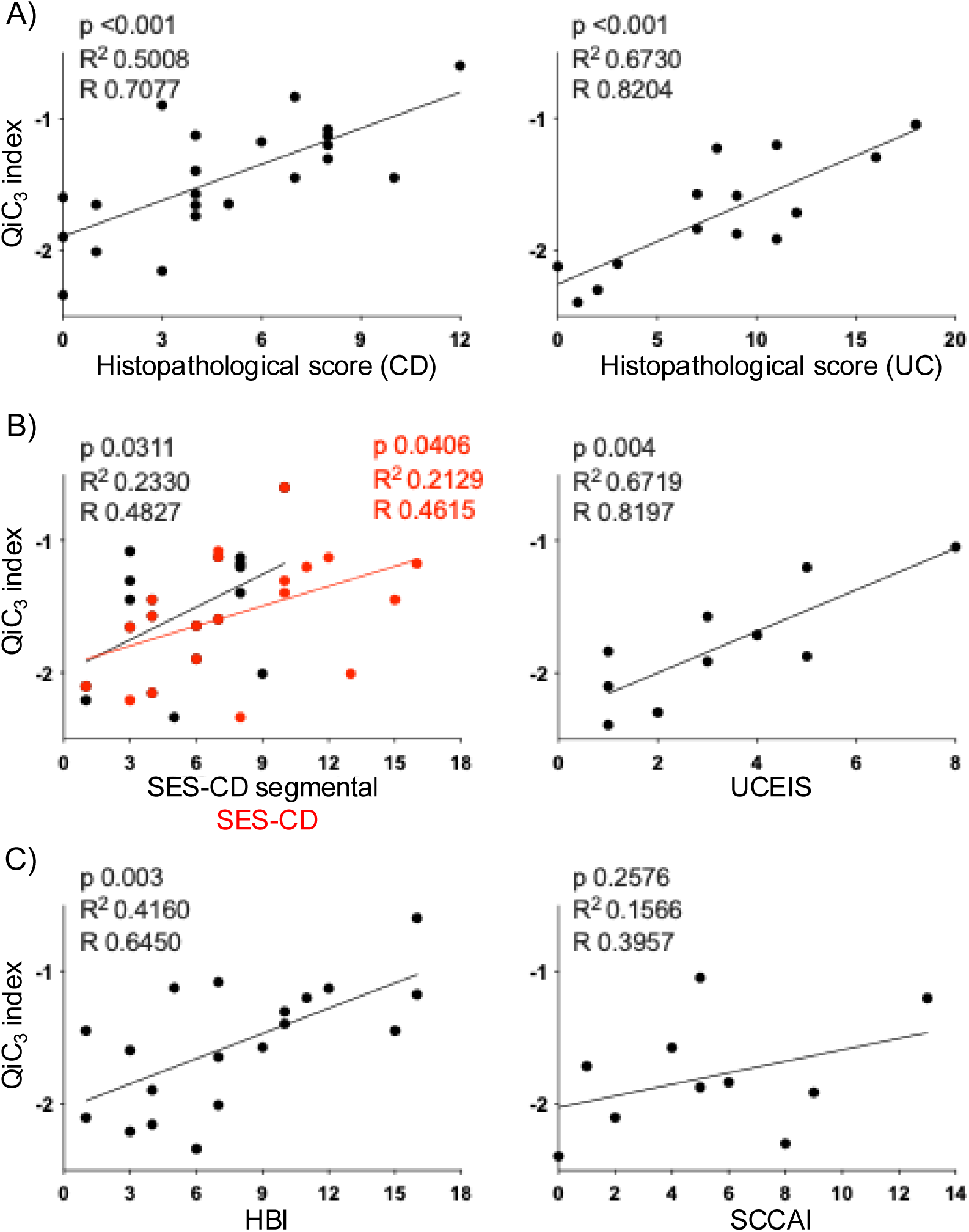
The QiC_3_ index reflects the histological and endoscopic status of the intestinal mucosa. Correlation of (A) histological scores, (B) endoscopic scores, or (C) symptom-based scores, with the QiC_3_ index in patients with CD or UC. For CD, endoscopic correlations include both total SES-CD and the segmental SES-CD scores corresponding to the most inflamed intestinal segment. These previously described and established histopathological, endoscopic, and symptom-based scoring systems are specific for either CD or UC. Spearman correlation test; SES-CD, Simple Endoscopic Score for Crohn’s Disease; UCEIS, Ulcerative Colitis Endoscopic Index of Severity; HBI, Harvey-Bradshaw Index (symptom-based score for CD); SCCAI, Simple Clinical Colitis Activity Index (symptom-based score for UC).

For a proportion of CD patients we measured the level of calprotectin in the feces the day before the mucosal biopsy was collected. We compared the QiC_3_ value of the most inflamed biopsy, as defined by histopathological evaluation, from each patient with the calprotectin level. The results showed a correlation between the QiC_3_ index and the fecal calprotectin levels (Supplementary Figure 5A). We also performed a correlation analysis to investigate a potential association of the number of epithelial or LP neutrophils with the level of fecal calprotectin in these CD patients. We found that fecal calprotectin correlated well with the number of LP neutrophils, and even better with the number of epithelial neutrophils (Supplementary Figure 5B).

### Results from a small validation cohort, and analysis of the responsiveness of the QiC_3_ index

To investigate the reproducibility of the new QiC_3_ index, we analyzed biopsies from a new group of IBD patients that all had endoscopically active disease (small validation cohort, n=8). We calculated and scored these samples using the QiC_3_ index. Again, the results showed an excellent separation from the control subjects (Figure 6A). Next, we set out to examine the responsiveness of the QiC_3_ index by evaluating the mucosal improvements induced by anti-TNFα treatment. Biopsies were collected from the endoscopically most inflamed segment of 10 CD patients with moderate to severe disease (HBI score of ≥7) before and after 12 weeks of adalimumab treatment. After treatment, patients were subdivided into those who reached remission and those who did not, as defined by four different parameters, respectively. The number of patients that achieved remission after 12 weeks of adalimumab treatment were 4 as defined by the histopathological score (GHAS ≤2); 5 as defined by the endoscopic score (SES-CD ≤2); and 8 as defined by either symptom levels (HBI score ≤4) or fecal calprotectin levels (≤200 µg/g). We then plotted the QiC_3_ scores before and after treatment for each of the groups (Figure 6B). The QiC_3_ index showed good conformity with the histopathological definition of remission, whereas the congruence with remission defined endoscopically, clinically, or by fecal calprotectin was somewhat poorer (Figure 6B). The results show that the QiC_3_ index displays good responsiveness to disease improvement, especially when disease activity is defined by the histological degree of activity (Figure 6B).

**Figure 6.**
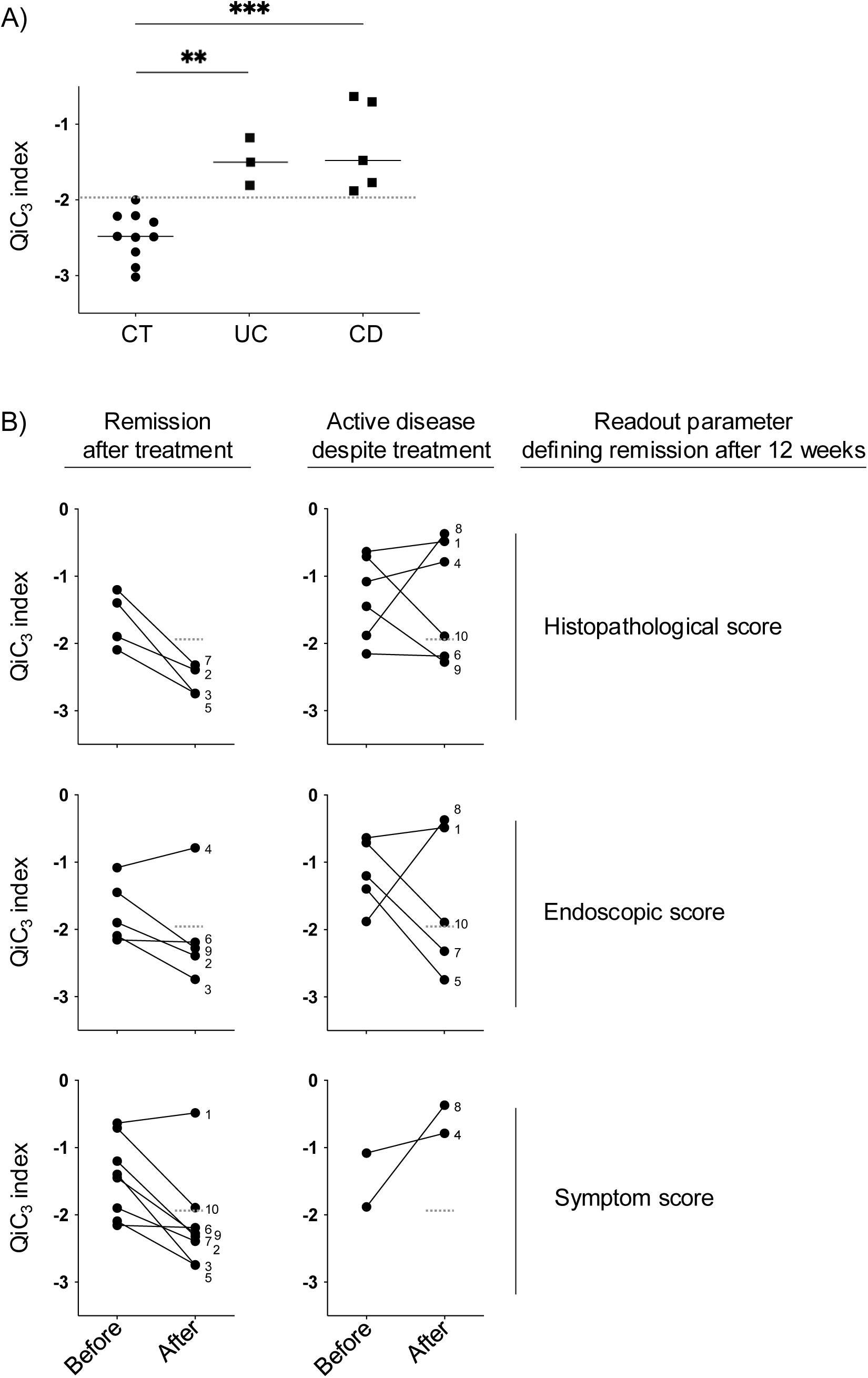
In a small validation cohort of IBD patients (UC n=3, CD=5), the QiC_3_ index scores showed an equally good separation between biopsies from IBD patients and control subjects as the previous dataset. Furthermore, the QiC_3_ index showed good responsiveness to disease improvement induced by anti-TNFα therapy. (A) The QiC_3_ index was tested on a small validation group of intestinal biopsies from UC (n=3) and CD (n=5) patients with endoscopically active disease. Lines show the median. (B) QiC_3_ index before and 12 weeks after starting adalimumab treatment (anti-TNFα therapy) in CD. Remission was defined histologically as GHAS ≤2, endoscopically as SES-CD score ≤2, symptomatically as HBI score ≤4, and by fecal calprotectin level as ≤200 µg/g. The patients defined as being in remission after 12 weeks according to fecal calprotectin levels and HBI scores, respectively, happened to be the same 8 patients, thus shown in the same graph.

### The QiC_3_ index values showed a significant correlation with the tissue expression levels of proinflammatory mediators

Inflammation is promoted by inflammatory cytokines such as IL-1β and TNFα, as well as other proinflammatory mediators such as S100A8 and S100A9.^25^ To investigate the association of disease activity as defined by the QiC_3_ index with the expression levels of proinflammatory mediators, we performed a correlation analysis between the QiC_3_ index values and S100A8, S100A9, IL-1β, and TNFα gene expression levels. For gene expression analysis, the intestinal samples were subjected to the Affymetrix Human Gene 2.1 ST microarray and then the normalized values were used for the correlation analysis. We found that the QiC_3_ index correlated significantly with the expression levels of S100A8, S100A9, IL-1β, and TNFα, respectively, in a mixture of inflamed and uninflamed samples from patients with CD (Figure 7).

**Figure 7.**
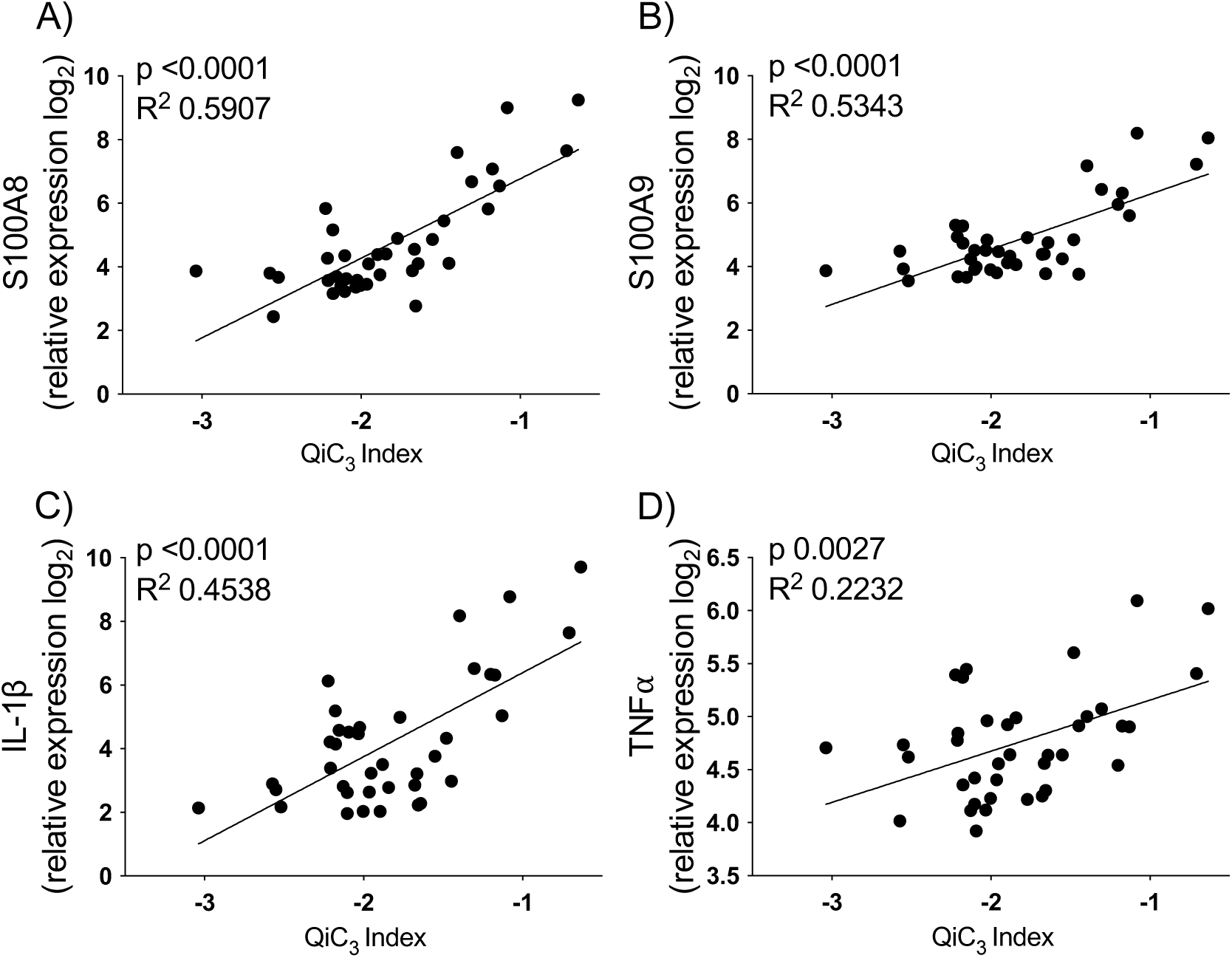
The QiC_3_ index showed significant correlation with the expression levels of proinflammatory mediators. Correlation of the gene expression level of (A) S100A8, (B) S100A9, (C) IL-1β, and (D) TNFα with the QiC_3_ index in patients with CD. The intestinal samples were subjected to the Affymetrix Human Gene 2.1 ST Array and gene expression values were extracted and normalized, and presented as Log_2_ values. Pearson correlation test.

## Discussion

Accurate quantitative assessment of histological disease activity has several potential applications in both clinical management and preclinical research in the field of IBD. The European Crohn’s and Colitis Organisation has stated that it is desirable and important that pathologists give an indication of the histological disease activity in their reports on mucosal tissue samples from IBD patients.^26,27^ However, the traditional histopathological examination of IBD biopsies is insufficient with regards to describing the degree of inflammation and it involves several highly person-dependent and subjective components.^2,3,28^ In this study, we developed an objective and responsive immunohistological index based on the numbers of various mucosal immune cells. This computerized and automated analysis of IBD biopsies holds the potential of giving a reliable and high-resolution measure of the inflammatory activity, thus suitable for monitoring of treatment responses and disease activity over time in clinical practice. In addition, it could be used as a standard tool in exploratory research and as an end-point tool in clinical trials evaluating new IBD pharmaceuticals. Finally, it would unburden work overloaded pathologists and shorten the response time-lapse. A standard histological scoring system would also make it possible to perform meta-analyses of several independent studies. In response to the increasing need for a quantitative and standardized tool for evaluation of intestinal samples, researchers have made efforts to develop automated histological assessment methods even in animal models of IBD.^29,30^ Recently, similar approaches have been reported for human biopsies.^31^ Expert gastrointestinal pathologists have primarily focused on defining qualitative and diagnostic criteria for IBD, and less on quantitative measures of the degree of inflammation in patients with an established diagnosis of IBD. Nevertheless, several semi-quantitative histopathological indices for UC and CD have indeed been proposed,^10–12,18,19^ but it has been difficult to agree on a common system for grading histological disease activity in IBD.^2,3^

Several studies have highlighted the importance of immune cells in the pathogenesis of IBD.^13^ Moreover, the infiltration of immune cells into the mucosal tissue is one of the central factors in histopathological assessments of intestinal IBD samples.^11,12^ In this study we examined three subsets of immune cells (i.e. neutrophils, macrophages and T cells) in two microanatomical mucosal compartments (the epithelium and the LP) to construct a new immunohistological index. We selected these subsets of immune cells to study since it is well-known that neutrophils accumulate in the intestinal mucosa of IBD patients,^32^ since the frequency of proinflammatory macrophages has been shown to be elevated in the inflamed mucosa of IBD patients,^33,34^ and since there are strong indications that mucosa-infiltrating proinflammatory T cells drive disease activity in IBD.^1^ Our results showed a significant increase in the numbers of epithelial and LP neutrophils, as well as epithelial macrophages, but not T cells,^35,36^ in mucosal samples from IBD patients compared to control subjects. Intestinal macrophages are traditionally considered to be situated subepithelially, primarily in the subepithelial zone directly underlying the epithelial cells, but not within the epithelium.^37,38^ Therefore it was surprising that the results showed significant CD68 positivity in the epithelial compartment, suggesting an increase in intraepithelially located macrophages. Importantly, biopsies from control subjects displayed much lower intraepithelial CD68 signal compared to biopsies from IBD patients. Analysis of the subepithelial zone did not show any differences in CD68 positivity, thus the increased epithelial CD68 positivity is a true epithelial signal and not an artifact produced by misinterpreted subepithelial macrophages. It is possible that macrophages enter the epithelium in the context of IBD and inflammation. Alternatively, the intraepithelial CD68 signal comes from protrusions from subepithelially located macrophages.^38–40^ Although unusual, it is known that CD68 can be expressed by other celltypes than macrophages, e.g. small populations of DCs, and it is thus theoretically possible that the intraepithelial CD68 signal represents some other celltype, either within the epithelium or with protrusions through the epithelium.^41,42^

Numerically, we used the logarithmic values of either summed or multiplied numbers of epithelial neutrophils, epithelial macrophages, and LP neutrophils. Using ROC analysis for investigating the ability of the indices to differ between IBD and control samples, we found that summing the three parameters gave a better separation than when they were multiplied. The QiC_3_ index could separate biopsies from active IBD patients and biopsies from control subjects with very high accuracy. In addition, the QiC3 index generated statistically significant separations between IBD biopsies with active and non-active disease, in both CD and UC. Moreover, we showed that the QiC_3_ index is equally suitable for colonic as for ileal biopsies, making it a global ileocolonic disease activity index. It has been suggested that there are differences in the numbers of T cells in the colon as compared with the small bowel,^1^ but our data showed that the numbers of T cells as well as the numbers of neutrophils and macrophages in uninflamed tissues were similar in ileal and colonic biopsies.

We validated the QiC_3_ index against previously described histopathological indices and found that QiC_3_ correlated strongly with established CD and UC indices.^6,9,43–45^ An advantage of the QiC_3_ index is that it can be used for both CD and UC.^9,10,46^ Although it is not possible to subclassify IBD patients based solely on histopathological examination, historically there has not been any common histopathological index for CD and UC.^3^ However, just recently a new index with such properties has been suggested.^9,10^ Several histopathological features, for example those describing architectural changes, are subjective and characterized by low interobserver as well as intraobserver reproducibility, making many previously described histopathological indices unreliable.^47^ In contrast, the QiC_3_ index, being a computerized, objective, and automated index, circumvents problems with reproducibility and interobserver variation. To explore whether there is potential to simplify the QiC_3_ index even further, we omitted the macrophage component and quantified the neutrophils collectively for the epithelium and the LP. Intriguingly, this simplified version of the index performed almost equally well as the full version. Indeed, several recent reports have highlighted the ability of neutrophil numbers to separate between inactive and active disease.^7,8,10,31,48,49^

In the field of IBD, it is well recognized that histopathological, endoscopic, and symptombased indices correlate poorly with each other.^3,50^ Corroborating these insights, our results showed that the QiC_3_ index correlated strongly with histopathological scores in both CD and UC. However, the QiC_3_ index showed a moderate to strong correlation with endoscopic evaluation too. The endoscopic scoring system SES-CD for CD evaluates four variables in the ileum, the right colon, the transverse colon, the left colon, and the rectum, generating a single score for the whole colon together with the terminal ileum; whereas the endoscopic scoring system UCEIS for UC is designed to evaluate a single segment.^20,21^ In addition, the SES-CD includes scoring of potential strictures, which may or may not be reflecting ongoing inflammation. These issues may explain why endoscopic evaluation correlated only weakly with the QiC_3_ index in patients with CD. As expected the correlation with fecal calprotectin was fairly good, but what was more interesting was that the correlation between the numbers of epithelial neutrophils with fecal calprotectin was even better. Correlation with the degree of symptoms was poor in UC but better in CD, which from clinical experience however would be expected to be the opposite.^5^ Finally, we found that the QiC_3_ index correlated significantly with the expression levels of proinflammatory mediators in patients with CD, possibly meaning that QiC_3_ index reflects the molecular inflammatory status of the gut mucosa.^51^ Several studies have described the phenomenon of existing microscopic inflammation being present in macroscopically uninflamed intestinal biopsies from UC and CD patients.^3,52–54^ There is also data to suggest that subclinical or microscopic inflammation is associated with an increased risk for clinical relapse, hospitalization, stricture formation, colorectal cancer, serious infection, and surgery.^3,52,53,55,56^ There is as yet no consensus on recommending histological remission as a treatment target in a general treat-to-target strategy, although it has been proposed.^7,8,51^ With the numerous new drugs under development, histological assessment of disease activity is being increasingly considered as an endpoint in clinical trials.^2–4^

In conclusion, we have introduced a new computerized, automated, objective, and responsive immunohistological index - the QiC_3_ index - which has the ability to assess histological disease activity in both CD and UC patients with high accuracy. The QiC_3_ index was constructed from the immunohistochemically defined numbers of epithelial neutrophils, epithelial macrophages, and the numbers of LP neutrophils. This new index is an objective tool that may be used in clinical management of IBD, in clinical trials for IBD, and in basic research in this field. Based on the components of the QiC3 index, we think that it also has potential to be applied in research on intestinal infectious diseases and research involving animal models for intestinal inflammation.

## Data Availability

All data produced in the present study are available upon reasonable request to the authors.

## Acknowledgements

The authors would like to thank Sven Almer for discussions and ethical approval; Medetect AB (Lund, Sweden) for kindly providing resources and immunohistochemical computerized technology; Britt-Marie Nilsson for technical assistance with tissue processing and serial sectioning; and finally, the staff at the endoscopy unit and the outpatient clinic at the Section for Gastroenterology at Skane University Hospital in Lund for supporting endoscopies, handling of human experimental material, and patient communication.

## Funding

This work was supported by the Hedlund Foundation, the Julin Foundation, the Bengt Ihre Stipend, the Swedish Society for Medical Research, the Royal Physiographic Society of Lund, Skane University Hospital donations, the Swedish Society of Medicine, an investigator-initiated study grant from AbbVie, research support from the Healthcare Region of Southern Sweden, and grants to researchers in public health care from the Swedish government (ALFSKANE-539811) to Jan Marsal.

## Disclosures

Erik Hertervig has served as consultant for Abbvie, Merck, Sharp & Dohme, and Takeda. Jonas S Erjefält is the founder and CEO of Medetect AB. Jan Marsal has served as a speaker, consultant or advisory board member for AbbVie, Amgen, Bayer, Biogen, BMS, Eli Lilly, Ferring, Galapagos/Alfasigma, Hospira, ITH, Janssen, Lument, MSD, Otsuka, Pfizer, Sandoz, Takeda, Tillotts and UCB; and has received grant support (not for this study) from AbbVie, BMS, Calpro AS, Carbiotix, Ferring, Fresenius Kabi, Pfizer, Svar Life Science and Takeda. Jan Marsal has received investigator-initiated study grants from AbbVie, Ferring, Pfizer, and Takeda. The other authors have no financial conflicts of interest.

## Author contributions

Mohammad Kadivar: study concept and design; performing experiments; analysis and interpretation of data; making of figures and tables; statistical analysis; writing and revision of manuscript. Manar Alyamani: analysis and interpretation of data; making of figures and tables; statistical analysis; writing and revision of manuscript. Michiko Mori: performing immunohistochemistry experiments; revision of manuscript. Maryam Kadivar: histopathological analysis and scoring; revision of manuscript. Jimmie Jönsson: performing image analysis, making of figures. Erik Hertervig: providing human experimental material; study concept and design; revision of manuscript. Olof Grip: providing human experimental material; study concept and design; revision of manuscript. Lena Svensson: interpretation of data; revision of manuscript; study supervision. Jonas Erjefält: analysis and interpretation of immunohistochemistry data; providing computerized immunohistochemistry reading methodology and materials; revision of manuscript. Jan Marsal: study concept and design; obtained ethical approval; analysis and interpretation of data; writing and revision of manuscript; recruitment of study subjects and providing human experimental material; study supervision; obtained funding.

## Abbreviations

CD: Crohn’s disease
UC: ulcerative colitis
GHAS: global histological activity scoring
HBI: Harvey–Bradshaw index
cGS: continuous Geboes histological score for UC
IEL: intraepithelial lymphocyte
LP: lamina propria
MPO: myeloperoxidase
ns: non significant
SES-CD: simple endoscopic activity score for Crohn’s disease
UCEIS: ulcerative colitis endoscopic index of severity
TNF: tumor necrosis factor

## Supplementary Figures

**Figure S1.** Two-color staining of intestinal tissues to define the epithelial layer and immune cells. Epithelial cells were stained for cytokeratins and β-catenin. Dark brown staining represents MPO, CD68 and CD3 immunoreactivity in control subjects, CD, and UC samples as shown.

**Figure S2.** ROC analysis of the numbers of neutrophils, macrophages, and T cells, in the LP, the epithelium, and the subepithelial zone, respectively. ROC curves show the sensitivity and specificity of neutrophils (A), macrophages (B), and T cells (C), in the LP, the epithelium, and the subepithelial zone, respectively, for discriminating UC and CD samples, respectively, from control subjects. Accuracy was measured by AUC. Accuracy was measured by calculations of area under curve (AUC); an AUC of 1.0 represents perfect discrimination.

**Figure S3.** Correlation analysis of the number of LP, epithelial, and subepithelial zone neutrophils, macrophages, and T cells, respectively, with each other and histopathological scores. (A) Spearman correlations between numbers of LP, epithelial, and subepithelial zone MPO^+^, CD68^+^ and CD3^+^ cells, respectively, with each other. Numbers represent P values. p<0.05 numbers are shown in red. (B) Spearman correlations between LP, epithelial, and subepithelial zone MPO^+^, CD68^+^ and CD3^+^ cells, respectively, and CD and UC histopathological scores. Numbers represent P values. p<0.05 numbers are shown in red. Parameters that comprise the QiC_3_ index are shown in light blue squares.

**Figure S4.** The numbers of neutrophils in the LP and the epithelium calculated collectively (MPO index), were significantly higher in inflamed intestinal tissues of IBD patients, as defined by histopathological indices, compared to control subjects. Graphs show biopsies from control subjects and biopsies from CD and UC patients with active (Act.) and non-active disease (Non-act.). Each dot represents an individual biopsy and bars are mean±SD. Student t-test; ns, non significant, **P<0.001, ****P<0.0001. ROC curves show the sensitivity and specificity of the collective mucosal neutrophil numbers for discriminating UC and CD biopsies with active inflammation from biopsies of control subjects. Accuracy was measured by calculations of area under curve (AUC); an AUC of 1.0 represents perfect discrimination. Optimal cutoffs were determined using Youden’s J statistic on ROC curves, with a cutoff of −1.97.

**Figure S5.** Correlation analysis of the QiC3 index and number of LP and epithelial neutrophils, respectively, with fecal calprotectin levels. Spearman correlation analysis of the QiC3 index (A), LP neutrophils (B), and epithelial neutrophils (C) with fecal-calprotectin. Optimal cutoffs were determined using Youden’s J statistic on ROC curves, with a cutoff of −1.96 for the QiC_3_ index.

## Notes

### Competing Interest Statement

Erik Hertervig has served as consultant for Abbvie, Merck, Sharp & Dohme, and Takeda. Jonas S Erjefalt is the founder and CEO of Medetect AB. Jan Marsal has served as consultant for AbbVie, BMS, Ferring, Galapagos, Lilly, Takeda, Tillotts. Jan Marsal has received investigator-initiated study grants from AbbVie, Ferring, Pfizer, and Takeda. The other authors have no financial conflicts of interest.

### Author Declarations

The study was performed in accordance with the declaration of Helsinki and was approved by the regional ethics committees in Lund and Linkoping, Sweden (Dnr. 2011/60 and 2011-201-31, respectively). Written informed consent was obtained from all subjects before they were included.

## References

1. Marsal J, Agace W. Targeting T-cell migration in inflammatory bowel disease. Journal of internal medicine. 2012;272(5):411–429.

2. Bryant R, Winer S, Travis S, Riddell R. Systematic review: histological remission in inflammatory bowel disease. Is ‘complete’remission the new treatment paradigm? An IOIBD initiative. Journal of Crohn’s and Colitis. 2014;8(12):1582–1597.

3. Walsh AJ, Bryant RV, Travis SP. Current best practice for disease activity assessment in IBD. Nature Reviews Gastroenterology & Hepatology. 2016;13(10):567–579.

4. Levesque BG, Sandborn WJ, Ruel J, Feagan BG, Sands BE, Colombel J-F. Converging goals of treatment of inflammatory bowel disease from clinical trials and practice. Gastroenterology. 2015;148(1):37–51. e31.

5. Turner D, Ricciuto A, Lewis A, et al. STRIDE-II: An Update on the Selecting Therapeutic Targets in Inflammatory Bowel Disease (STRIDE) Initiative of the International Organization for the Study of IBD (IOIBD): Determining Therapeutic Goals for Treat-to-Target strategies in IBD. Gastroenterology. 2021;160(5):1570–1583.

6. Neri B, Mossa M, Scucchi L, Sena G, Palmieri G, Biancone L. Histological scores in inflammatory bowel disease. J Dig Dis. 2021;22(1):9–22.

7. Pai RK, Hartman DJ, Rivers CR, et al. Complete Resolution of Mucosal Neutrophils Associates With Improved Long-Term Clinical Outcomes of Patients With Ulcerative Colitis. Clin Gastroenterol Hepatol. 2020;18(11):2510–2517 e2515.

8. Pai RK, Jairath V, Feagan BG. Editorial: histologic normalisation in ulcerative colitis. Aliment Pharmacol Ther. 2020;51(3):399–401.

9. Fumery M, Chatelain D. Histological Scores in Inflammatory Bowel Disease: A New Kid in the Block. J Crohns Colitis. 2021;15(10):1603–1604.

10. Lang-Schwarz C, Angeloni M, Agaimy A, et al. Validation of the ‘Inflammatory Bowel Disease-Distribution, Chronicity, Activity [IBD-DCA] Score’ for Ulcerative Colitis and Crohn s Disease. J Crohns Colitis. 2021;15(10):1621–1630.

11. D’haens GR, Geboes K, Peeters M, Baert F, Penninckx F, Rutgeerts P. Early lesions of recurrent Crohn’s disease caused by infusion of intestinal contents in excluded ileum. Gastroenterology. 1998;114(2):262–267.

12. Geboes K, Riddell R, Öst A, Jensfelt B, Persson T, Löfberg R. A reproducible grading scale for histological assessment of inflammation in ulcerative colitis. Gut. 2000;47(3):404–409.

13. McBride RB, Suarez-Farinas M, Ko HM, Chen X, Liu Q, Harpaz N. Density of Biopsy Sampling Required to Ensure Accurate Histological Assessment of Inflammation in Active Ulcerative Colitis. Inflamm Bowel Dis. 2023;29(11):1706–1712.

14. Magro F, Lopes J, Borralho P, et al. Comparing the Continuous Geboes Score With the Robarts Histopathology Index: Definitions of Histological Remission and Response and their Relation to Faecal Calprotectin Levels. J Crohns Colitis. 2020;14(2):169–175.

15. Mosli MH, Feagan BG, Sandborn WJ, et al. Histologic evaluation of ulcerative colitis: a systematic review of disease activity indices. Inflamm Bowel Dis. 2014;20(3):564–575.

16. Vespa E, D’Amico F, Sollai M, et al. Histological Scores in Patients with Inflammatory Bowel Diseases: The State of the Art. J Clin Med. 2022;11(4).

17. Zenlea T, Yee EU, Rosenberg L, et al. Histology Grade Is Independently Associated With Relapse Risk in Patients With Ulcerative Colitis in Clinical Remission: A Prospective Study. Am J Gastroenterol. 2016;111(5):685–690.

18. Mosli MH, Feagan BG, Zou G, et al. Development and validation of a histological index for UC. Gut. 2017;66(1):50–58.

19. Marchal-Bressenot A, Salleron J, Boulagnon-Rombi C, et al. Development and validation of the Nancy histological index for UC. Gut. 2017;66(1):43–49.

20. Daperno M, D’Haens G, Van Assche G, et al. Development and validation of a new, simplified endoscopic activity score for Crohn’s disease: the SES-CD. Gastrointestinal endoscopy. 2004;60(4):505–512.

21. Travis SP, Schnell D, Krzeski P, et al. Developing an instrument to assess the endoscopic severity of ulcerative colitis: the Ulcerative Colitis Endoscopic Index of Severity (UCEIS). Gut. 2012;61(4):535–542.

22. Bergqvist A, Andersson CK, Mori M, Walls AF, Bjermer L, Erjefält JS. Alveolar T-helper type-2 immunity in atopic asthma is associated with poor clinical control. Clinical Science. 2015;128(1):47–56.

23. Mori M, Andersson CK, Svedberg KA, et al. Appearance of remodelled and dendritic cell-rich alveolar-lymphoid interfaces provides a structural basis for increased alveolar antigen uptake in chronic obstructive pulmonary disease. Thorax. 2013;68(6):521–531.

24. Bolstad BM, Irizarry RA, Åstrand M, Speed TP. A comparison of normalization methods for high density oligonucleotide array data based on variance and bias. Bioinformatics. 2003;19(2):185–193.

25. De Souza HS, Fiocchi C. Immunopathogenesis of IBD: current state of the art. Nature Reviews Gastroenterology & Hepatology. 2015.

26. Magro F, Langner C, Driessen A, et al. European consensus on the histopathology of inflammatory bowel disease. Journal of Crohn’s and Colitis. 2013;7(10):827–851.

27. Magro F, Doherty G, Peyrin-Biroulet L, et al. ECCO Position Paper: Harmonization of the Approach to Ulcerative Colitis Histopathology. J Crohns Colitis. 2020;14(11):1503–1511.

28. Mosli MH, Feagan BG, Zou G, et al. Reproducibility of histological assessments of disease activity in UC. Gut. 2015;64(11):1765–1773.

29. Kozlowski C, Jeet S, Beyer J, et al. An entirely automated method to score DSS-induced colitis in mice by digital image analysis of pathology slides. Disease models & mechanisms. 2013;6(3):855–865.

30. Rogers R, Eastham-Anderson J, DeVoss J, et al. Image Analysis-Based Approaches for Scoring Mouse Models of Colitis. Veterinary pathology. 2016;53(1):200–210.

31. Gui X, Bazarova A, Del Amor R, et al. PICaSSO Histologic Remission Index (PHRI) in ulcerative colitis: development of a novel simplified histological score for monitoring mucosal healing and predicting clinical outcomes and its applicability in an artificial intelligence system. Gut. 2022;71(5):889–898.

32. Davies JM, Abreu MT. The innate immune system and inflammatory bowel disease. Scandinavian journal of gastroenterology. 2015;50(1):24–33.

33. Bain C, Scott C, Uronen-Hansson H, et al. Resident and pro-inflammatory macrophages in the colon represent alternative context-dependent fates of the same Ly6Chi monocyte precursors. Mucosal immunology. 2013;6(3):498–510.

34. Thiesen S, Janciauskiene S, Uronen-Hansson H, et al. CD14hiHLA-DRdim macrophages, with a resemblance to classical blood monocytes, dominate inflamed mucosa in Crohn’s disease. Journal of leukocyte biology. 2014;95(3):531–541.

35. Royset ES, Sahlin Pettersen HP, Xu W, et al. Deep learning-based image analysis reveals significant differences in the number and distribution of mucosal CD3 and gammadelta T cells between Crohn’s disease and ulcerative colitis. J Pathol Clin Res. 2023;9(1):18–31.

36. Kondo A, Ma S, Lee MYY, et al. Highly Multiplexed Image Analysis of Intestinal Tissue Sections in Patients With Inflammatory Bowel Disease. Gastroenterology. 2021;161(6):1940–1952.

37. Hegarty LM, Jones GR, Bain CC. Macrophages in intestinal homeostasis and inflammatory bowel disease. Nat Rev Gastroenterol Hepatol. 2023.

38. Joeris T, Muller-Luda K, Agace WW, Mowat AM. Diversity and functions of intestinal mononuclear phagocytes. Mucosal Immunol. 2017;10(4):845–864.

39. Schulz O, Jaensson E, Persson EK, et al. Intestinal CD103+, but not CX3CR1+, antigen sampling cells migrate in lymph and serve classical dendritic cell functions. J Exp Med. 2009;206(13):3101–3114.

40. Chikina AS, Nadalin F, Maurin M, et al. Macrophages Maintain Epithelium Integrity by Limiting Fungal Product Absorption. Cell. 2020;183(2):411–428 e416.

41. Farache J, Koren I, Milo I, et al. Luminal bacteria recruit CD103+ dendritic cells into the intestinal epithelium to sample bacterial antigens for presentation. Immunity. 2013;38(3):581–595.

42. Niess JH, Brand S, Gu X, et al. CX3CR1-mediated dendritic cell access to the intestinal lumen and bacterial clearance. Science. 2005;307(5707):254–258.

43. Park J, Kang SJ, Yoon H, et al. Histologic Evaluation Using the Robarts Histopathology Index in Patients With Ulcerative Colitis in Deep Remission and the Association of Histologic Remission With Risk of Relapse. Inflamm Bowel Dis. 2022;28(11):1709–1716.

44. Peyrin-Biroulet L, Arenson E, Rubin DT, et al. A comparative evaluation of the measurement properties of three histological indices of mucosal healing in ulcerative colitis: Geboes Score, Robarts Histopathology Index, and Nancy Index. J Crohns Colitis. 2023.

45. Villanacci V, Antonelli E, Lanzarotto F, Bozzola A, Cadei M, Bassotti G. Usefulness of Different Pathological Scores to Assess Healing of the Mucosa in Inflammatory Bowel Diseases: A Real Life Study. Sci Rep. 2017;7(1):6839.

46. Villanacci V, Del Sordo R, Parigi TL, Leoncini G, Bassotti G. Inflammatory Bowel Diseases: Does One Histological Score Fit All? Diagnostics (Basel). 2023;13(12).

47. Bressenot A, Salleron J, Bastien C, Danese S, Boulagnon-Rombi C, Peyrin-Biroulet L. Comparing histological activity indexes in UC. Gut. 2014:gutjnl-2014-307477.

48. Ohara J, Maeda Y, Ogata N, et al. Automated Neutrophil Quantification and Histological Score Estimation in Ulcerative Colitis. Clin Gastroenterol Hepatol. 2025;23(5):846–854 e847.

49. Parigi TL, Cannatelli R, Nardone OM, et al. Neutrophil-only Histological Assessment of Ulcerative Colitis Correlates with Endoscopic Activity and Predicts Long-term Outcomes in a Multicentre Study. J Crohns Colitis. 2023;17(12):1931–1938.

50. Lemmens B, Arijs I, Van Assche G, et al. Correlation between the endoscopic and histologic score in assessing the activity of ulcerative colitis. Inflammatory bowel diseases. 2013;19(6):1194–1201.

51. Danese S, Roda G, Peyrin-Biroulet L. Evolving therapeutic goals in ulcerative colitis: towards disease clearance. Nat Rev Gastroenterol Hepatol. 2020;17(1):1–2.

52. Baars JE, Nuij VJ, Oldenburg B, Kuipers EJ, van der Woude CJ. Majority of patients with inflammatory bowel disease in clinical remission have mucosal inflammation. Inflammatory bowel diseases. 2012;18(9):1634–1640.

53. Bessissow T, Lemmens B, Ferrante M, et al. Prognostic value of serologic and histologic markers on clinical relapse in ulcerative colitis patients with mucosal healing. The American journal of gastroenterology. 2012;107(11):1684–1692.

54. Floren CH, Benoni C, Willen R. Histologic and colonoscopic assessment of disease extension in ulcerative colitis. Scand J Gastroenterol. 1987;22(4):459–462.

55. Bitton A, Peppercorn MA, Antonioli DA, et al. Clinical, biological, and histologic parameters as predictors of relapse in ulcerative colitis. Gastroenterology. 2001;120(1):13–20.

56. Mikolajczyk AE, Cohen NA, Watson S, et al. Assessment of the Degree of Variation of Histologic Inflammation in Ulcerative Colitis. Inflamm Bowel Dis. 2023;29(2):222–227.

