## Supplemental materials for "QiC_3_: A novel automated quantitative immunohistological disease activity index for ileocolonic Crohn’s disease and ulcerative colitis"

Figure S1

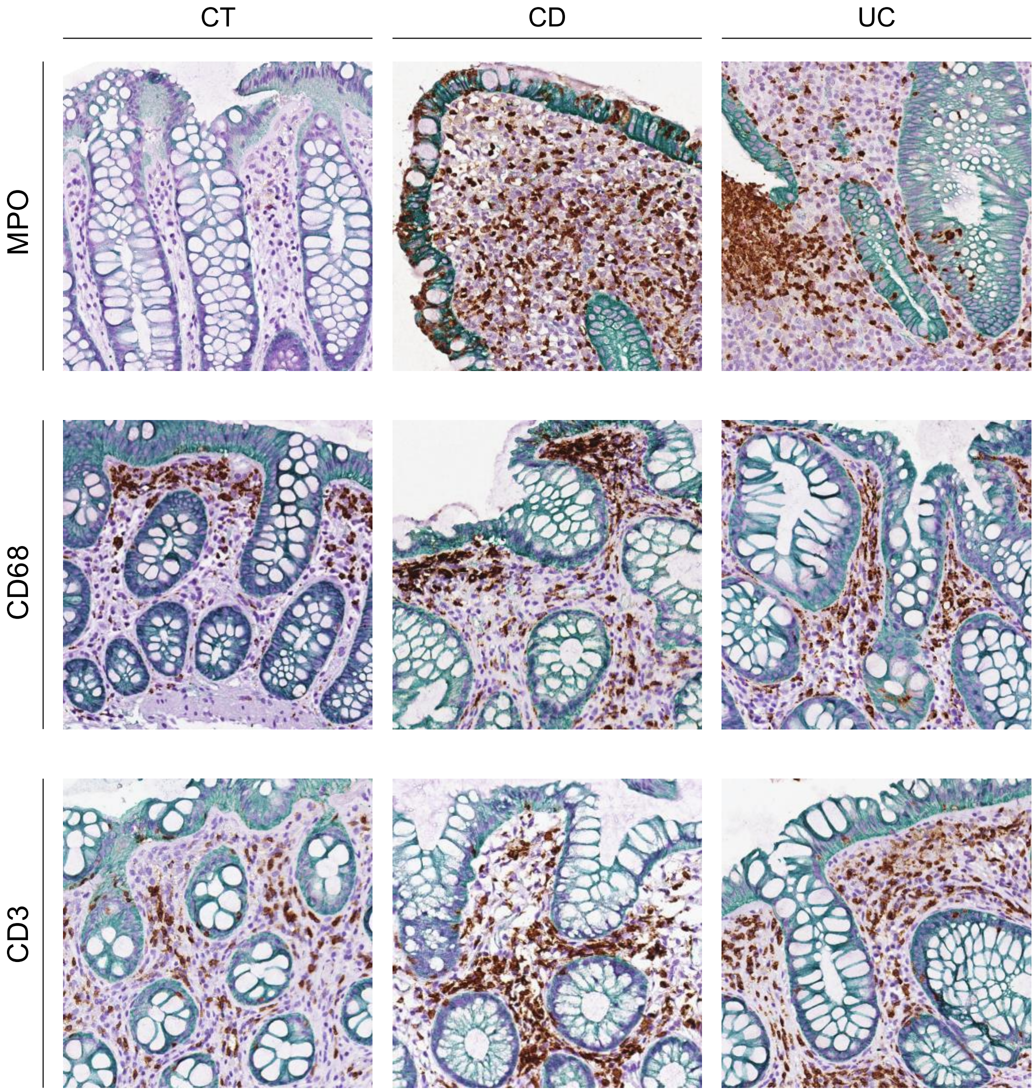

Figure S2

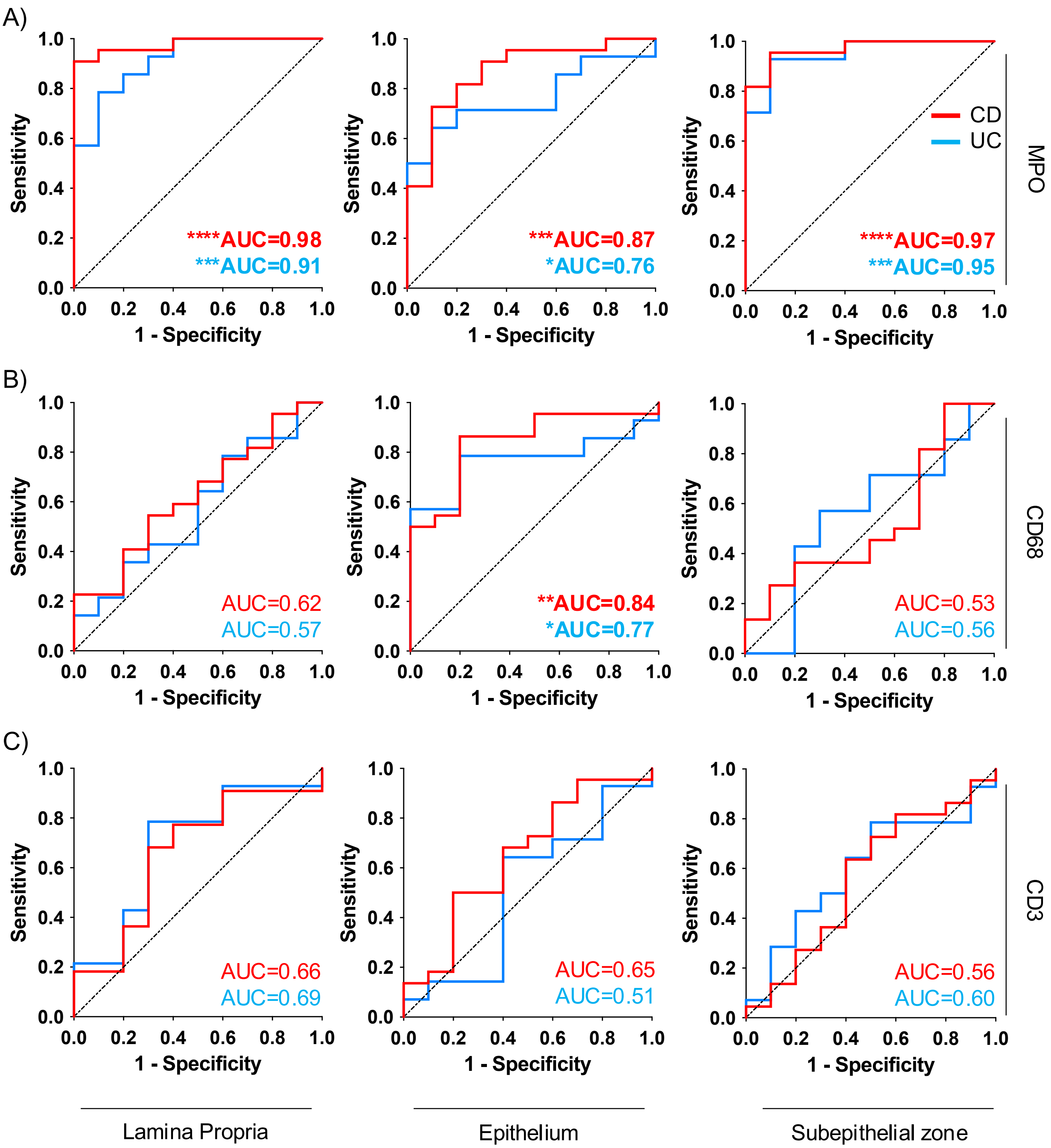
A)
B)
C)
Sensitivity
1 - Specificity
CD
UC
MPO
CD68
CD3
****AUC=0.98
***AUC=0.91
***AUC=0.87
*AUC=0.76
****AUC=0.97
***AUC=0.95
AUC=0.62
AUC=0.57
**AUC=0.84
*AUC=0.77
AUC=0.53
AUC=0.56
AUC=0.66
AUC=0.69
AUC=0.65
AUC=0.51
AUC=0.56
AUC=0.60
Lamina Propria
Epithelium
Subepithelial zone

Figure S3

A)

| | MPO epithelium | MPO LP | MPO subepithelium | CD68 epithelium | CD68 LP | CD68 subepithelium | CD3 epithelium | CD3 LP | CD3 subepithelium |
|---|---|---|---|---|---|---|---|---|---|
| MPO epithelium | | **<0.0001** | **<0.0001** | **<0.0001** | 0.9841 | 0.2027 | 0.6132 | 0.1167 | 0.2452 |
| MPO LP | | | **<0.0001** | **<0.0001** | 0.7364 | 0.6023 | 0.4961 | 0.4471 | 0.5197 |
| MPO subepithelium | | | | **<0.0001** | 0.6351 | 0.1913 | 0.6854 | 0.1677 | 0.3081 |
| CD68 epithelium | | | | | 0.7692 | 0.1024 | **0.0312** | 0.1294 | 0.2 |
| CD68 LP | | | | | | **<0.0001** | 0.5564 | 0.6346 | 0.0736 |
| CD68 subepithelium | | | | | | | **0.0273** | **0.0057** | **0.0013** |
| CD3 epithelium | | | | | | | | **0.0038** | **<0.0001** |
| CD3 LP | | | | | | | | | **<0.0001** |
| CD3 subepithelium | | | | | | | | | |

B)

| Histological score | MPO epithelium | MPO LP | MPO subepithelium | CD68 epithelium | CD68 LP | CD68 subepithelium | CD3 epithelium | CD3 LP | CD3 subepithelium |
|---|---|---|---|---|---|---|---|---|---|
| CD | **<0.0001** | **<0.0001** | **<0.0001** | **0.0027** | 0.9329 | 0.9305 | 0.1029 | 0.0678 | 0.3686 |
| UC | **<0.0001** | **<0.0001** | **<0.0001** | **<0.0001** | 0.8755 | **0.0409** | 0.2359 | **0.0193** | 0.1173 |

Figure S4

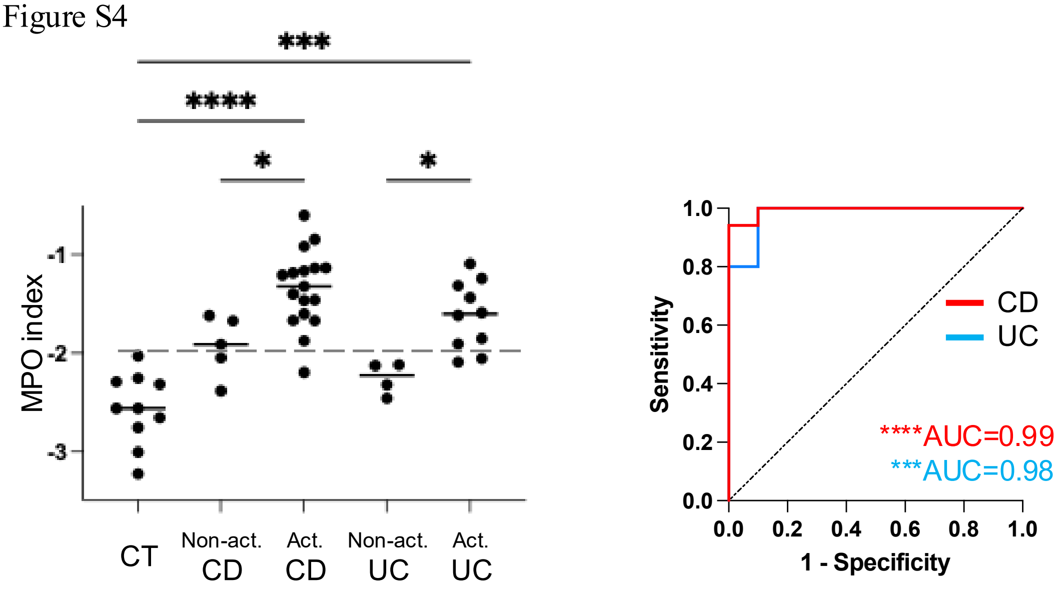

***
****
*
*
MPO index
-1
-2
-3
CT
Non-act.
CD
Act.
CD
Non-act.
UC
Act.
UC
1.0
0.8
0.6
0.4
0.2
0.0
Sensitivity
0.0
0.2
0.4
0.6
0.8
1.0
1 - Specificity
CD
UC
****AUC=0.99
***AUC=0.98

Figure S5

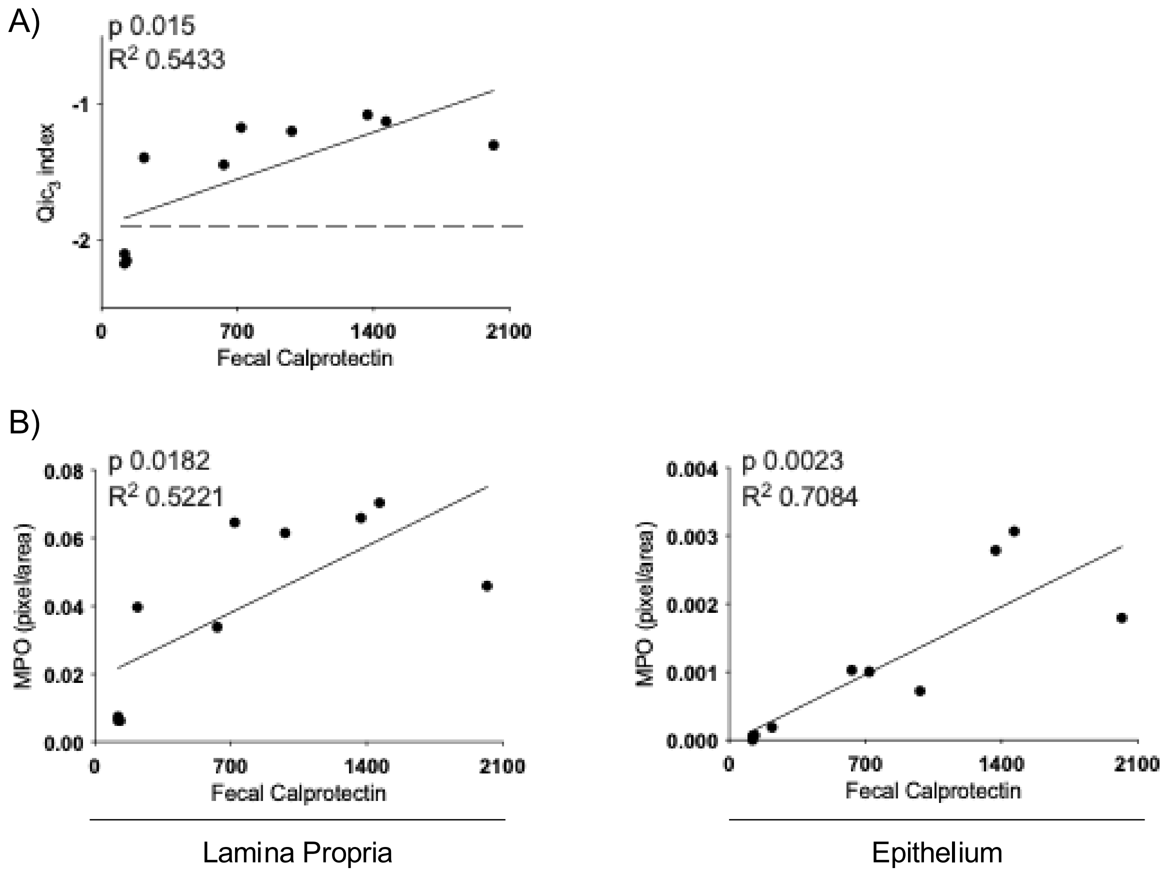
A)
p 0.015
R2 0.5433
Qic3 index
-1
-2
0
700
1400
2100
Fecal Calprotectin
B)
p 0.0182
R2 0.5221
MPO (pixel/area)
0.08
0.06
0.04
0.02
0.00
0
700
1400
2100
Fecal Calprotectin
Lamina Propria
p 0.0023
R2 0.7084
MPO (pixel/area)
0.004
0.003
0.002
0.001
0.000
0
700
1400
2100
Fecal Calprotectin
Epithelium
